# Hospitalization as reliable indicator of second wave COVID-19 pandemic in eight European countries

**DOI:** 10.1101/2020.12.31.20249084

**Authors:** Mattia Allieta, Davide Rossi Sebastiano

**Affiliations:** Ronin Institute Montclair, NJ 07043 USA; Neurophysiology Unit, IRCCS-Neurological Institute “Carlo Besta”, via Celoria 11, 20133, Milan, Italy

**Keywords:** COVID-19, SARS-CoV-2, Second Wave, Europe, ICU, hospitalization

## Abstract

Time dependent reproduction number (*R*_t_) is one of the most popular parameters to track the impact of COVID-19 pandemic. However, especially at the initial stages, R_t_ can be highly underestimated because of remarkable differences between the actual number of infected people and the daily incidence of people who are tested positive. Here, we present the analysis of daily cumulative number of hospitalized (HP) and intensive care unit (ICU) patients both in space and in time in the early phases of second wave COVID-19 pandemic across eight different European countries, namely Austria, Belgium, Czech Republic, France, Italy, Portugal, Spain, and United Kingdom. We derive simple model equations to fit the time dependence of these two variables where exponential behavior is observed. Growth rate constants of HP and ICU are listed, providing country-specific parameters able to estimate the burden of SARS-COV-2 infection before extensive containment measures take place. Our quantitative parameters, fully related to hospitalizations, are disentangled from the capacity range of the screening campaign, for example the number of swabs, and they cannot be directly biased by the actual number of infected people. This approach can give an array of reliable indicators which can be used by governments and healthcare systems to monitor the dynamics of COVID-19 epidemic.

## 1. Introduction

Second wave COVID-19 pandemic constitutes an ongoing global threat and particularly in European Union (EU) which, according to World Health Organization (WHO), is again one of the epicenters of coronavirus epidemic (Han et al., 2020; Rypdal et al., 2020). From the first week of September, SARS-COV2 is rapidly spreading through almost all the EU countries and the cumulative incidence of official reported COVID-19 confirmed cases has reached more than 13 million on November 9^th^.

Usually, the time dependent reproduction number *R*_t_, (the average number of secondary cases generated by an infectious individual at time t) is considered one of the most important and informative parameters to track the epidemic trends (Riccardo et al., 2020; Rypdal et al., 2020). However, when it is huge the number of cases of a particular disease per unit of population (i.e. the morbility or morbidity rate, MB), as in COVID-19 pandemic, R_t_ can be underestimated, simply because the diagnostic possibilities of a healthcare system are overwhelmed and the difference between actually infected people and infected people who are tested positive is very high. In this context, the calculation of R_t_ is susceptible to mistakes which are very difficultly to be disentangled (Fan et al, 2020; Perico et al., 2020; Riccardo et al., 2020; Stefanelli et al., 2020; Whittaker et al., 2020).

On the other hand, mortality rates suffer from a time delay compared to the trend of the epidemic: the mean delay of 15 days between the death and the infection of COVID-19 makes this parameter virtually impractical for the decision-making, although it is one of the most reliable indicator of the pandemic by a theoretical point of view (García-Basteiro et al., 2020).

Even more than the epidemic trends as itself, from the beginning of COVID-19 pandemic, two key variables represent, the major problems which the health care systems must face with: the number of hospitalized patients (HP) and the number of Intensity Care Units (ICU) patients. In each EU country, HP and ICU are affected by the limited available resources, a dramatic bottleneck with respect to the possibility of an adequate care of each patient affected by COVID-19. Hence, because of R_t_ is an index strictly depending on the ratio between the number of swabs performed and MB and, at the same time, the mortality rate is a parameter temporally too far from the real situation to be used in the challenging epidemic context. HP and ICU are probably the only parameters representing a good compromise between reliability and solidity besides being remarkably useful in the decision making related to the COVID-19 pandemic.

Mathematical modeling of spreading of respiratory infectious diseases is a well-established field in epidemiology (Wallinga and Teunis, 2004) and, recently, many works have been devoted to model SARS-COV2 spreading and the virus transmission dynamics (as example, for Italy, see Riccardo et al., 2020, Allieta et al., 2020). Moreover, these valuable theoretical efforts, statistical analysis obtained by parametrizing the ongoing epidemiological data could provide reliable starting experimental parameters to perform more accurate predictive calculations.

In this work, we focused on the relationship between the number of HP and ICU by deriving a simple model equation able to fit the observed time evolution of cumulative incidence for eight selected EU countries, namely Austria, Czech Republic, Italy, France, Belgium, Portugal, Spain, United Kingdom (UK), at the start of second wave COVID-19 pandemic, from August to November 2020. This allows us to obtain consistent parameters to strictly monitor the local dynamics of COVID-19 epidemic in all the country considered.

## 2. Materials and Methods

### 2.1 Demographic and Epidemiological data

The official demographic data of the resident population updated on January 1^st^, 2020 for each European Union country selected were taken from Eurostat. We collected data of COVID-19 epidemic for the eight European countries (i.e. Austria, Belgium, Czech Republic, France, Italy, Portugal, Spain and United Kingdom) from official websites where data are aggregated at national levels and published in form of dashboard, daily or periodical reported. All the data used in this work are accessible and have been publicly published (see also Table S1, supporting information, for all the details on demographic and epidemiological data sources). The data of COVID-19 pandemic were collected from August 1^st^ to October 31^st^ for all the country considered except Spain where data were available only from August 20^th^ to October 31^st^.

### 2.2 Derivation of the time evolution equation of daily cumulative incidence of the number of hospitalized and intensive care unit patients related to COVID-19

We assume that HP at time *t* is N’_HP_ and, over a short time interval of duration Δ*t* from *t* to (*t*+Δ*t*), HP evolves to *k*_HP_Δ*t*N’_HP_ for some constant *k*_HP_ defined as growth rate constant of HP. We can then approximate the change of population size N’_HP_(t+Δ) – N’_HP_(*t*) to *k*_HP_Δ*t*N’_HP_(t) according to the following equality:

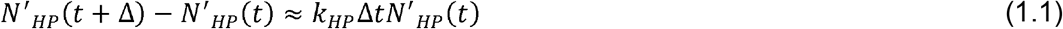

Dividing both side by Δt gives:

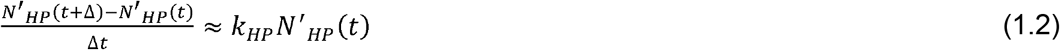

The duration of Δt must be short enough to ensure that the population size does not change too much and by considering the limit of Δ→ 0 under the assumption of differentiable N’_HP_(t) function, it follows:

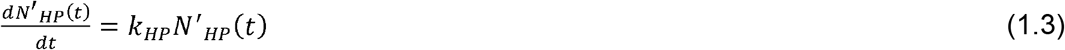

We can solve equation (1.3) using separation of variables and by imposing that *N*′_*HP*_(*t*) = *N*_*HP*_ (*t*) - *N*_*HP,e*_ where *N*_*HP*_ (*t*) is the ongoing cumulative HP at time *t* and *N*_*HP,e*_ is the cumulative HP of the “environment”. More precisely, since the second wave develops from an equilibrium situation where *N*_*HP*_ is not null, we assume for simplicity that the number of HP tends to an asymptotic “environmental” value exhibited before the N′_HP_(*t*) started to grow exponentially.

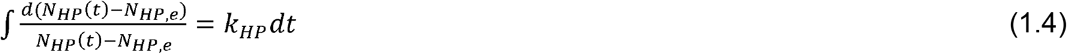

Hence,

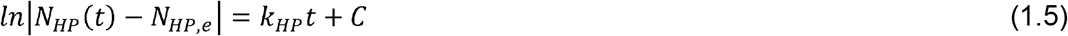

Solving for N_HP_ (t) we get the following exponential growth equation:

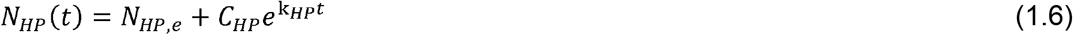

We applied the same assumptions to derive the differential equation related to the time evolution of ICU which, by implementing the above conditions, reads as:

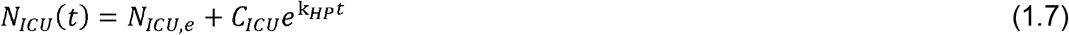

Since it is reasonable to consider that *N*_ICU_(t) can be strictly related to N_HP_(t) such that a function like *N*_ICU_ = *N*_ICU_[*N*_HP_(t),t] is more appropriate to describe its evolution. However, our goal is to analyze *N*_ICU_(t), *N*_HP_(t) separately to define an empirical relationship between them. In this context, we used equations (1.5), (1.6) to fit the time evolution of *N*_ICU_, *N*_HP_ and to refine values of environmental hospitalized and ICU constants (*N*_HP,e_, *N*_ICU_,_e_), empirical constants (*C*_HP_, *C*_ICU_) and exponential growth rate parameters (*k*_HP_, *k*_ICU_) directly against the observed data. Unit of measurement of *k*_HP_, *k*_ICU_ are in reciprocal day (d^-1^) which allows to define generation time constants *G*_HP_ = 1/*k*_HP_, *G*_ICU_=1/*k*_ICU_ as the time in days (d) which elapsed between initial population and the population at *t* of HP and ICU, respectively.

Furthermore, we defined a rate of conversion between N_HP_ and N_ICU_ patients as *ICU rate of hospitalization patients* (*R*_*ICU*_) and we made a connection between variables by adopting the simple equation:

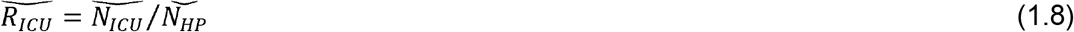

where 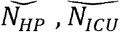 are expressed as cumulative incidence per 100000 inhabitants.

## 3. Results

Results are summarized In figures S1-S4 (supporting information), where we reported the cumulative incidence as number of total HP and ICU patients for all the countries.

All the profiles were fitted by using equations (1.6), (1.7) and the results from non-linear squares regression are listed in Table S2. Models are in good agreement with both observed HP and ICU data sets as testified by high goodness of fit, i.e. coefficient of determination R^2^, ranging from 0.956 to 0.997 and from 0.962 and 0.996 for HP and ICU, respectively.

We noted that the lowest (k_HP_ = 0.056) and highest (*k*_HP_ =0.115) values belong to neighboring Portugal and Spain, respectively, followed by France (*k*_HP_ =0.082) and Belgium (*k*_HP_ =0.084). We found that the 37.5 % of investigated countries display a *k*_HP_ ≈ 0.06 with an average *k*_HP_ of 0.075 ±0.018, according to values distribution (figure 1a). When considering ICU parameters, we reported that lowest value is belong again to Portugal (k_ICU_ = 0.049) while highest ones to Belgium (*k*_ICU_ =0.090) and Spain (*k*_ICU_ =0.086). The values distribution is similar to HP (figure 1a) with the 37.5 % of countries showing *k*_*ICU*_ ≈ 0.04 and an average *k*_*ICU*_ of 0.059 ± 0.019. *G*_HP_ varied from 9 to 18 days and *G*_ICU_ from 11 to 26 days.

**Figure 1.**
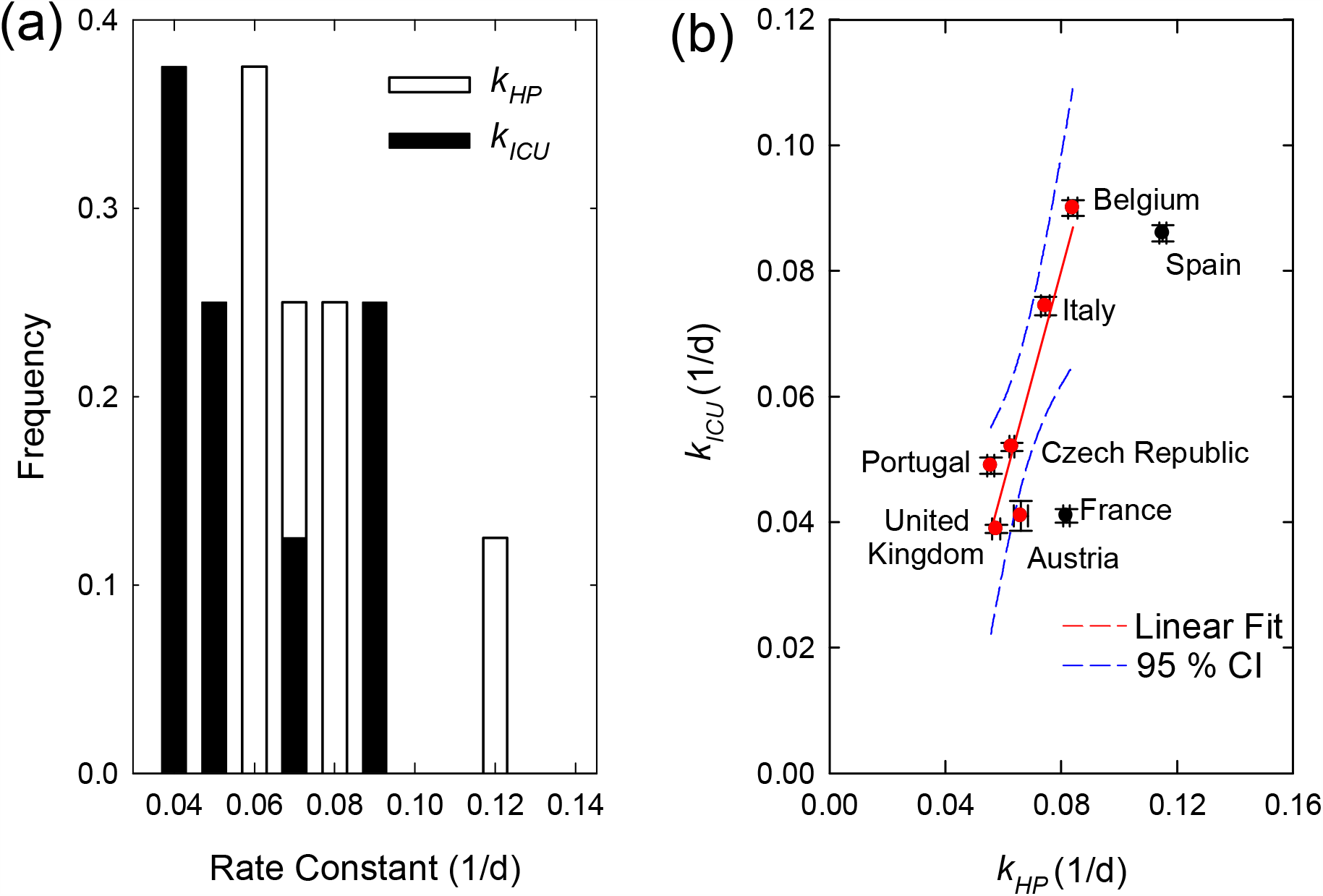
Correlation between Cumulative Incidence HP and Cumulative incidence ICU. (a) Observed distribution of rate constants HP (*k*_HP_) and ICU (*k*_ICU_) determined in each region. (b) Correlation between *k*_HP,_ *k*_ICU ;_ red and black dots represent countries in which *k*_ICU_/ *k*_HP_ is included or not included in the CI=95% of the linear fit model, respectively.

Surprisingly, we found that average *k*_*HP*_ and *k*_*ICU*_ are comparable and they can be related between each other as shown in Fig.1(b). A linear trend between the constants is then confirmed for all the countries, excluding France and Spain(figure 1b). This paves the way to delineate a more general correlation where for a given amount of HP a given amount of ICU can be then associated.

In figure 2a, we present the universal correlation between N_HP_ and N_ICU_ obtained by replotting all the observed data according to equation (1.8). R_ICU_ obtained for all the countries together with R^2^ determination coefficient are listed in Table S2 and ranked in figure 2b.

**Figure 2.**
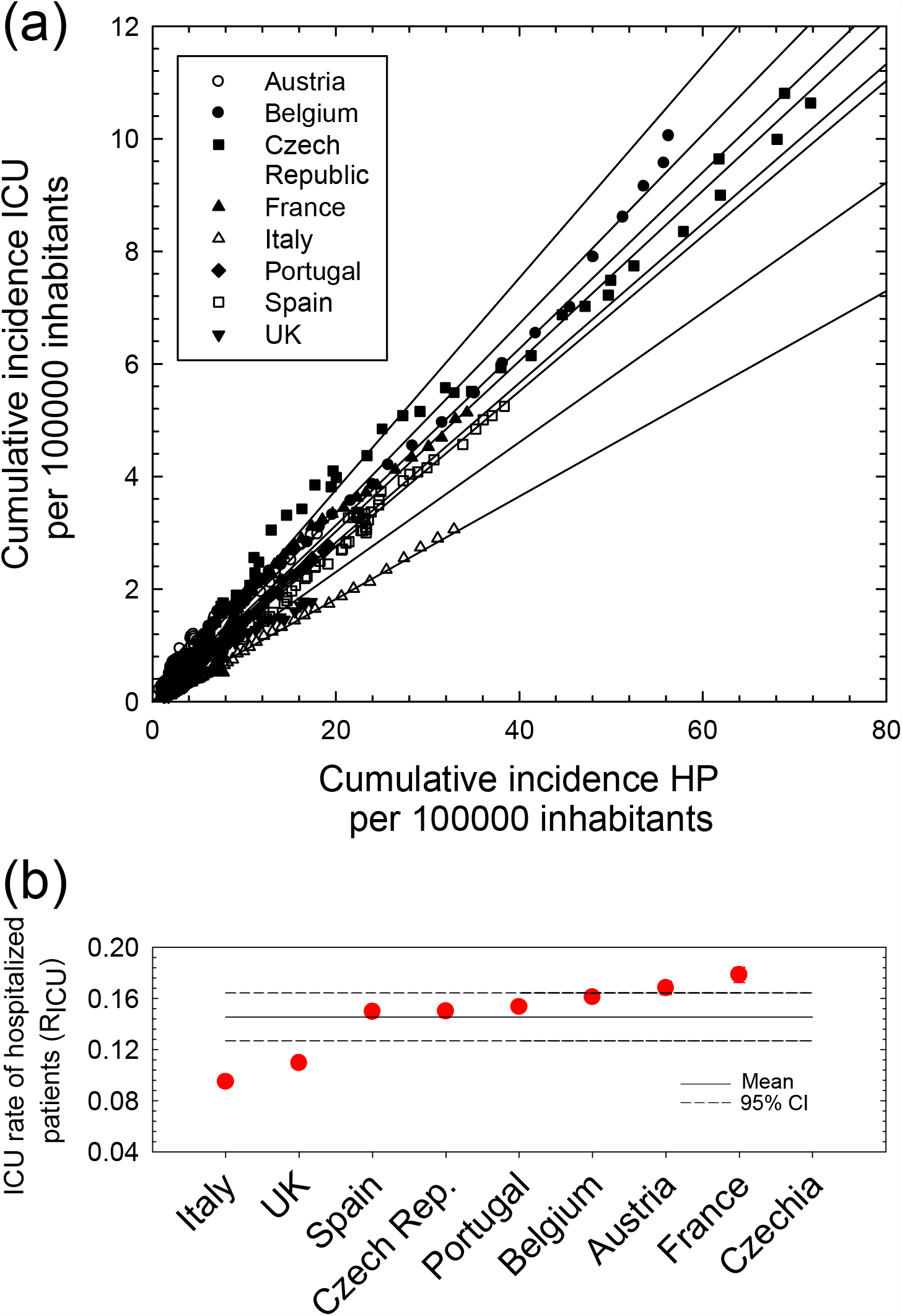
Correlation between Cumulative Incidence HP and Cumulative incidence ICU. (a) Correlation between cumulative incidence HP and Cumulative incidence ICU for each country analyzed. Data are normalized over Country Population. Solid lines are linear fitting model. (b) ICU rate of hospitalized patients (R_ICU_) compared and ranked for all the selected countries; red and black dots represent countries in which R_ICU_ is included or not included in the CI=95% of the linear fit model, respectively.

For the most of the countries analyzed, R_ICU_ followed the relation (1.8), excluding Czech Republic and France where higher R_ICU_ is observed when the cumulative incidence HP is below 200000/100000 Inhabitants. We reported that 5 countries display an R_ICU_ (%) close to the mean value 14.6% (95% CI: 12.7%, 16.4%) while Italy and UK exhibit R_ICU_ ≈ 10 %. The highest R_ICU_ ≈18 % is shown by France.

## 4. Discussion

In the year 2020, the COVID-19 pandemic determined the health and economic policy in all the EU countries. Based on the Chinese experience in the Hubei province and in the absence of effective treatments against the virus, to date, the most effective measures of public health to contain the pandemic have been non-pharmaceutical interventions such as quarantine, social distancing, and isolation of infected (Anderson et al., 2020). Although the restrictive measures taken by EU governments are necessary for the containment of the pandemic, they caused and they are still causing huge difficulties at individual, social and economic levels. Hence, given the impossibility of maintaining these restrictive measures for an unlimited period, all governments have worked to achieve a satisfactory balance based on reconciling economic and health needs. This can be done by restricting or by loosening the constraints imposed by the different epidemiological scenarios depending on the so called COVID-19 warning levels. In this context, it seems to be essential to have reliable and “manageable” indices to monitor the evolution of the epidemic trends. From a theoretical point of view, the actual reproduction number of infected people and the average reproduction number of secondary cases generated by an infectious individual (R_t_) could be considered one of the most reliable and informative parameters to use. However, a huge difficulty is represented by the large number of infected people in an asymptomatic or mild condition (Oran and Topol, 2020). The lack of large-scale diagnostic tests (swabs) mainly due to economic and logistic reasons adopted by the different governmental authorities could cause a serious underestimation of the actual number of COVID-19 cases in the population (Perico et al., 2020; Riccardo et al., 2020; Whittaker et al., 2020), namely in case of high-incidence of the epidemic (Fan et al, 2020; Stefanelli et al., 2020). The problem of under-reporting the total number of actual infected patients distorts the epidemic trends (Fernández-Fontelo et al., 2020), making government measures potentially ineffective or even out of time. On the other hand, mortality rates suffer from a time delay compared to the trend of the epidemic. The mean delay of 15-20 days between the contraction of COVID-19 infection and the death makes this parameter virtually impractical for decision-making (García-Basteiro et al., 2020). Here, we considered N_HP_ and N_ICU_ taken as a cumulative incidence in eight EU countries (Austria, Belgium, Czech Republic, France, Italy, Portugal, Spain and United Kingdom) in the period between August 1^st^ and October 31^st^ (for Spain between August, 20^th^ and October 31^st^), evaluating their relationship by means of several indices to determine if some of them can be considered effective to monitor the pandemic trends. In particular, by considering August 1^st^ as a starting point, we believe that the exponential growth of cumulative incidence observed up to October 31^st^, could offer a wide time window to extract intrinsic characteristic parameters related to the SARS-COV2. Moreover, this can be done before the effects of more stringent containment measures start to deeply affect the time evolution of epidemic.

As preliminary considerations, first we want to point out that we did not consider the cumulative number of new COVID-19 tested positive cases (N_t_), for the reasons described previously, i.e. N_t_ is strictly dependent on the number of swabs performed and it can be biased by several factors like underreporting, delays in recording as well as errors in classification of cases. Then, we only focused on the data of the second wave of COVID- 19 epidemic because the beginning of the pandemic was quite different across the countries considered and the analysis techniques to monitor the spreading of the epidemic (mainly molecular swabs) were developed during the pandemic peak. Moreover, the last part of the summer period (August) can be considered as a sort of steady-state of the epidemic characterized by consistent epidemiological data which are useful to analyze the subsequent variations in the daily incidence of hospitalizations from COVID-19. Indeed, it should be noted that similar approaches were impossible to follow during the first wave where the “submerged” infections were too much to allow an accurate estimation of the actual numbers (Apolone et al., 2020).

Even if the magnitude of the cumulative incidence of HP and ICU is quite different from one country to another, we found that the fitted growth rate constants are similar between each other, confirming that HP and ICU are strictly related in linear time-independent relationship obviously until effective containment measures were taken. Interestingly, UE countries apparently unrelated shared the same evolution of the pandemic trends: after about forty days from August 1^st^, in which the epidemic had a low and / or linear growth rate, the growth rate constants related to HP and ICU for all the studied UE countries started to have an exponential regime as marked by vertical dashed lines in Fig.S1-S4). We observed Austria which started to show exponential behavior of *N*_HP_ after September 8^th^ followed by Portugal and Belgium on September 12^th^ and 13^th^, respectively. Czech Republic, France and United Kingdom shared the same onset on September 14^th^ while Italy featured late onset on October 7^th^. Again, Spain displayed a one-of-a-kind time dependence showing exponential growth chart only after October 13^th^. The origin of this relationship is not completely explainable for the huge socio-demographic, environmental/climatic factors, even if three main causes could have strongly favored the second wave at the beginning of autumn season: the reopening of schools (Larosa et al., 2020), the lowering of the temperatures (Guasp et al., 2020; Rovetta and Castaldo, 2020) and of the solar irradiance (Guasp et al., 2020). The analysis of such correlations is beyond the scope of this paper and it will be the subject of our future investigations, although it is possible to hypothesize that the southern countries (Italy and Spain) had fewer infections than northern ones due to the hot summer of 2020. Beyond the “absolute” numbers related to HP and ICU, an “intrinsic” COVID-19-related epidemic trends seemed to emerge, again until effective containment measures were taken.

On the other hand, other parameters seem to be specific to each individual country, mainly K_HP_ and K_ICU_. Since K_HP_ and K_ICU_ are related to the generation time (generation time = 1/K), these two indices can be profitably used to predict when hospital systems and / or intensive care units may become saturated. This could be important to promptly implement effective containment measures, as the resources available for each country are not infinite. It should be noted that the only limitation in the use of the parameters related to the hospitalization of COVID-19 patients is the delay of these indices with respect to the actual situation (Garcia-Basteiro et al., 2020). However, it has been reported that this delay is less pronounced than those relating to deaths caused by the virus (Garcia-Basteiro et al., 2020).

Finally, R_icu_ represents the “conversion rate” between HP and ICU (i.e., hospitalized patients who subsequently need to be transferred to intensive care units) and, together with N_icu_ could be used to (roughly) evaluate the efficiency of the first-level health care (home therapy and prompt hospitalization), albeit indirectly. N_icu_ depends more on the intrinsic dynamics of the COVID-19 epidemic (albeit conditioned by government measures and socio-demographic and environmental characteristics) than effectiveness of the health system. When both N_icu_ and R_icu_ are high, the ICU (the number of patients in intensive care units) depends on the large number of hospitalized patients according to the above linear relationship between HP and ICU assuming an adequate first-level health care. Vice versa, when N_icu_ is high and R_icu_ is low, the health system is probably inadequate and the high ICU can depend on the high number of patients admitted directly to intensive care, due to insufficient home therapy and / or delayed hospitalization.

## 5. Conclusion

Epidemiologically, the second wave COVID-19 pandemic is an intricate phenomenon, where some intrinsic characteristics of the transmission dynamics of the SARS-COV 2 are strictly entangled with socio-demographic and environmental variables which are not easy to interpret However, we showed that observed parameters like the number of hospitalization and ICU can be used as reliable indicators to map the ongoing epidemic evolution. This can provide a tool to plan adequately containment measures which can be taken in the light of forecasts adapted to the real situation.

## Data Availability

The data that support the findings were derived from the following resources available in the public domain:
https://www.ages.at
https://www.sciensano.be
https://www.mzcr.cz
https://www.gouvernement.fr
https://www.salute.gov.it
https://www.sns.gov.pt
https://www.mscbs.gob.es
https://www.gov.uk
https://ec.europa.eu/eurostat

https://www.ages.at

https://www.sciensano.be

https://www.mzcr.cz

https://www.gouvernement.fr

https://www.salute.gov.it

https://www.sns.gov.pt

https://www.mscbs.gob.es

https://www.gov.uk

https://ec.europa.eu/eurostat

## 6. Acknowledgements

It is impossible to name all the people who helped us in this difficult period, but we must thank the informal group “Legere”, made up of doctors and researchers from Bergamo, Brussels, Frankfurt am Main and Genoa, for their continuous support.

## Patients Involvement

No patients were involved

## SUPPORTING INFORMATION

**Figure S1.**
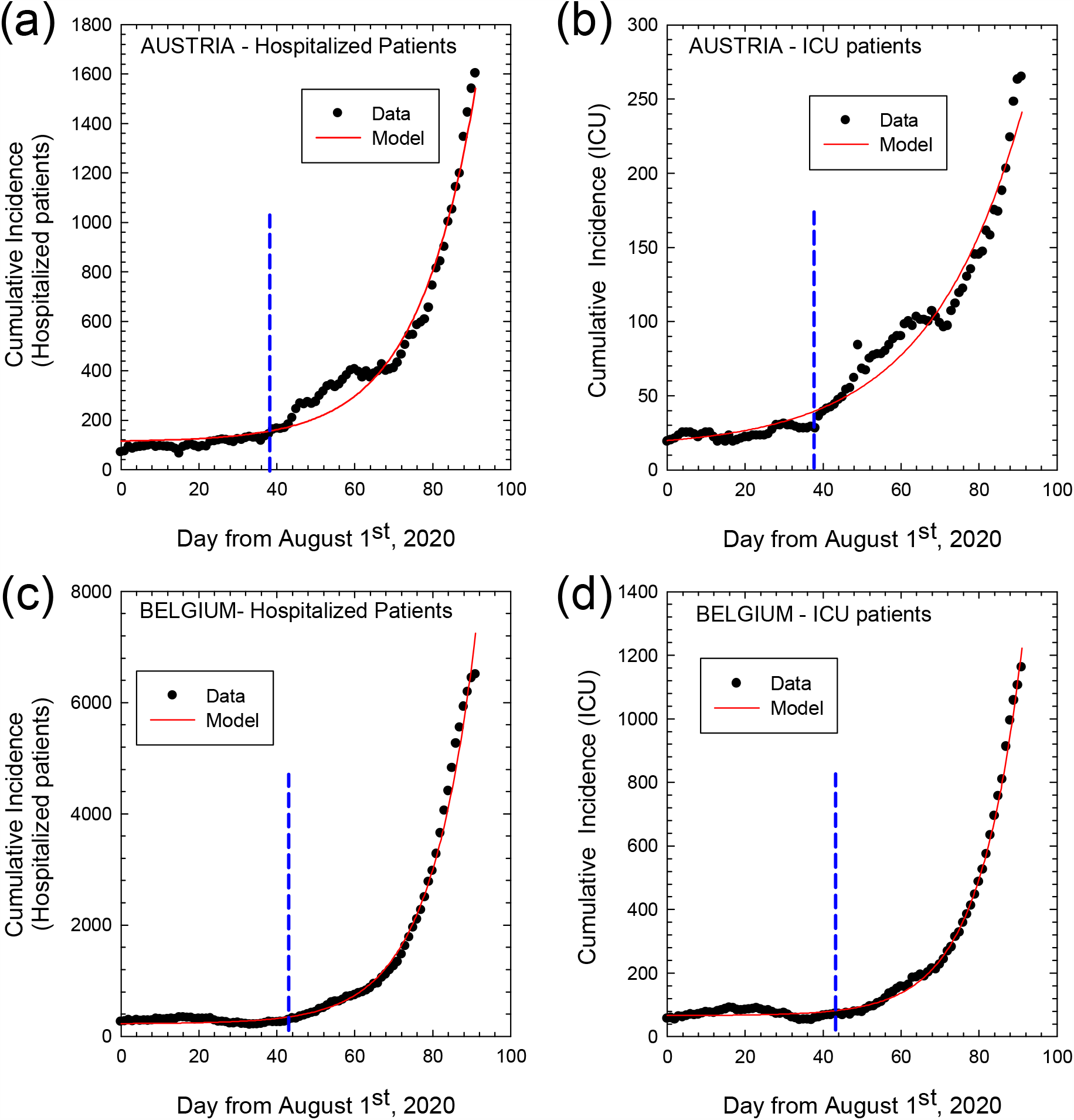
Cumulative Incidence for hospitalized and ICU patients. Cumulative incidence as numbers of total hospitalized patients (left panel) and ICU patients (right panel) from August 1^st^ to October 31^st^, 2020 for COVID-19 in Austria (a), (b) and Belgium (c), (d), respectively. Dots represent observed data and solid lines the fitting curves, respectively. Vertical dashed lines mark the day after which exponential regime is observed.

**Figure S2.**
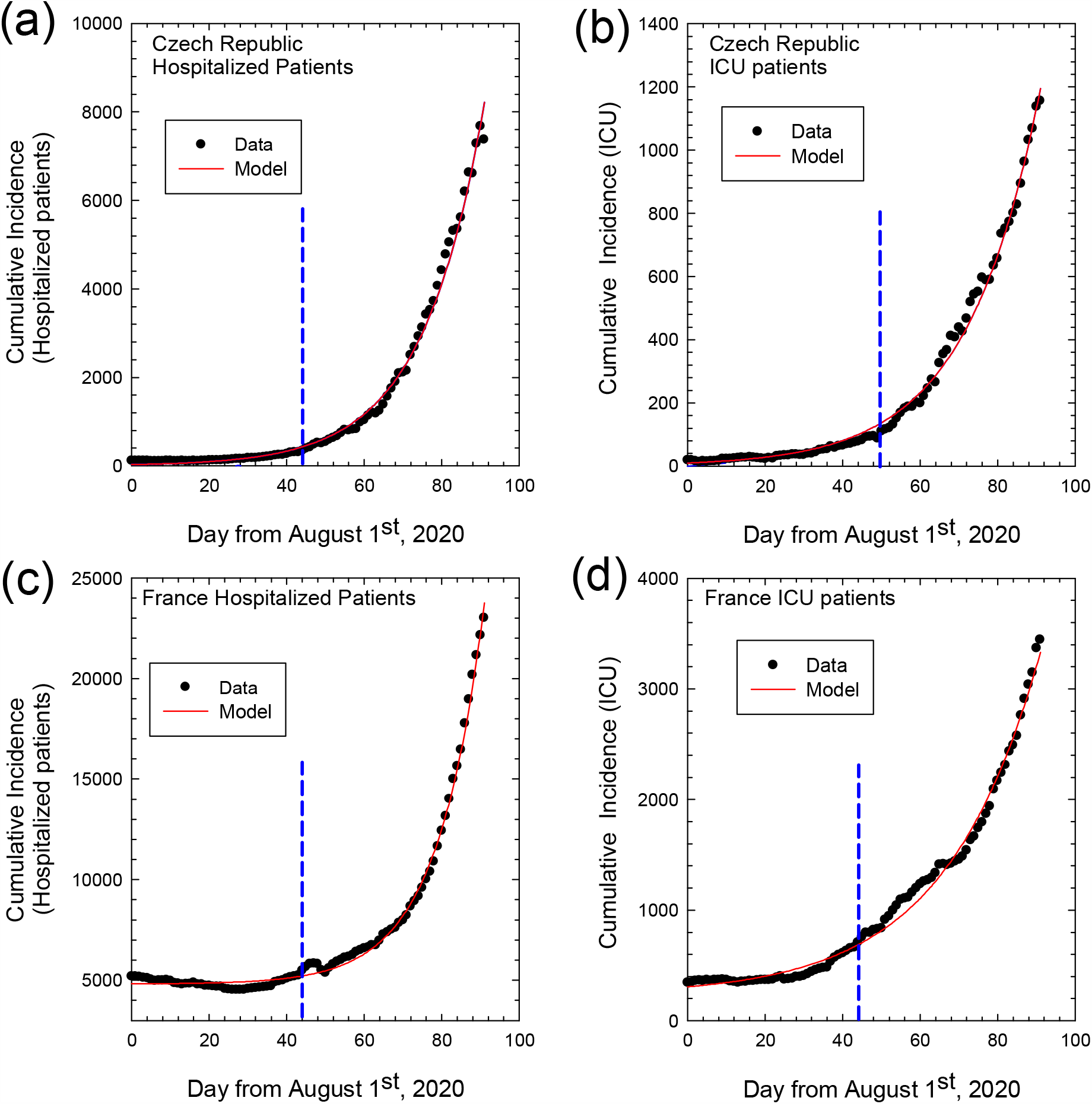
Cumulative Incidence for hospitalized and ICU patients. Cumulative incidence as numbers of total hospitalized patients (left panel) and ICU patients (right panel) from August 1^st^ to October 31^st^, 2020 for COVID-19 in Czech Republic (a), (b) and France (c), (d). Dots represent observed data and solid lines the fitting curves, respectively. Vertical dashed lines mark the day after which exponential regime is observed.

**Figure S3.**
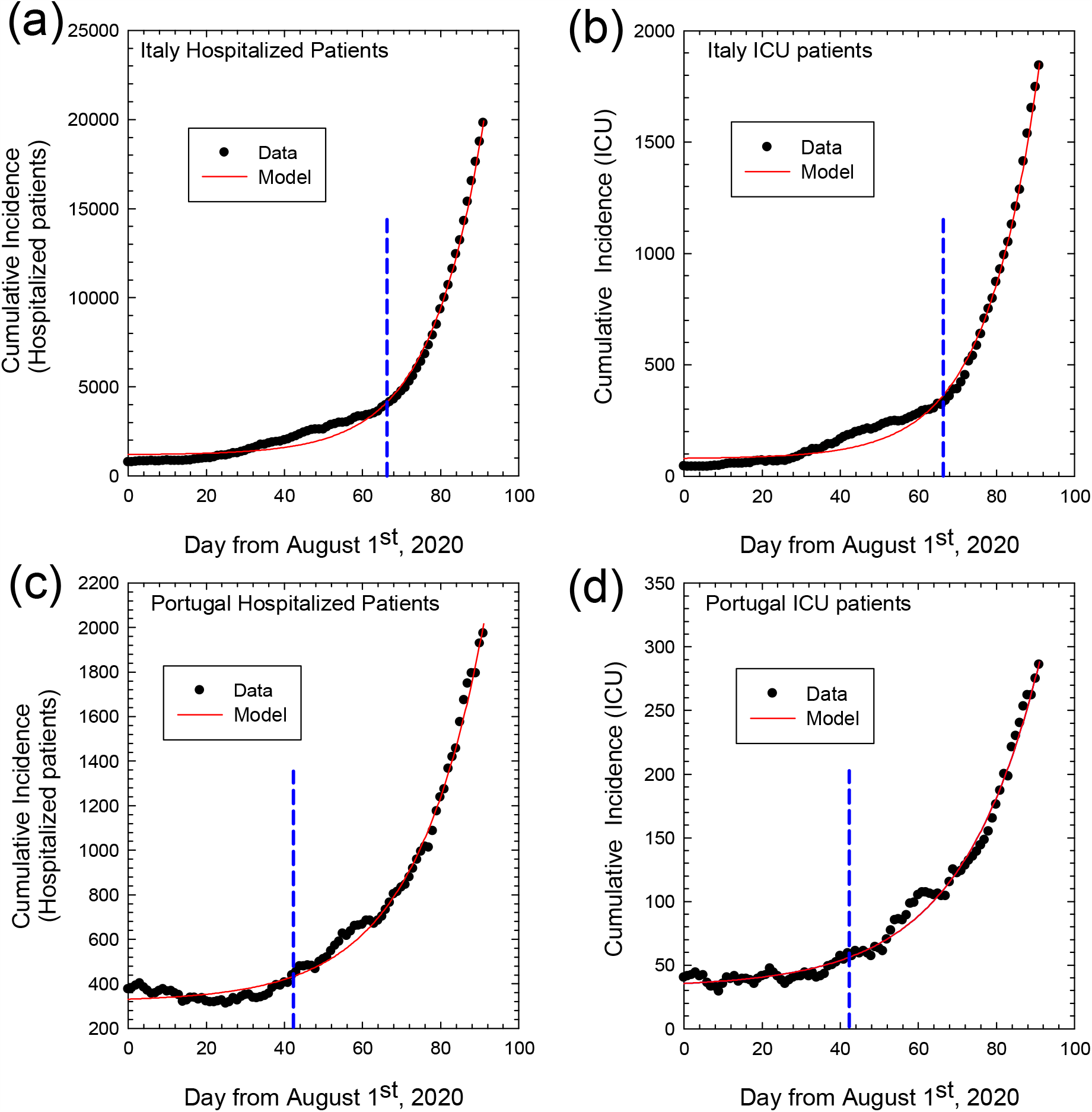
Cumulative Incidence for hospitalized and ICU patients. Cumulative incidence as numbers of total hospitalized patients (left panel) and ICU patients (right panel) from August 1^st^ to October 31^st^, 2020 for COVID-19 in Italy (a), (b) and Portugal (c), (d). Dots represent observed data and solid lines the fitting curves, respectively. Vertical dashed lines mark the day after which exponential regime is observed.

**Figure S4.**
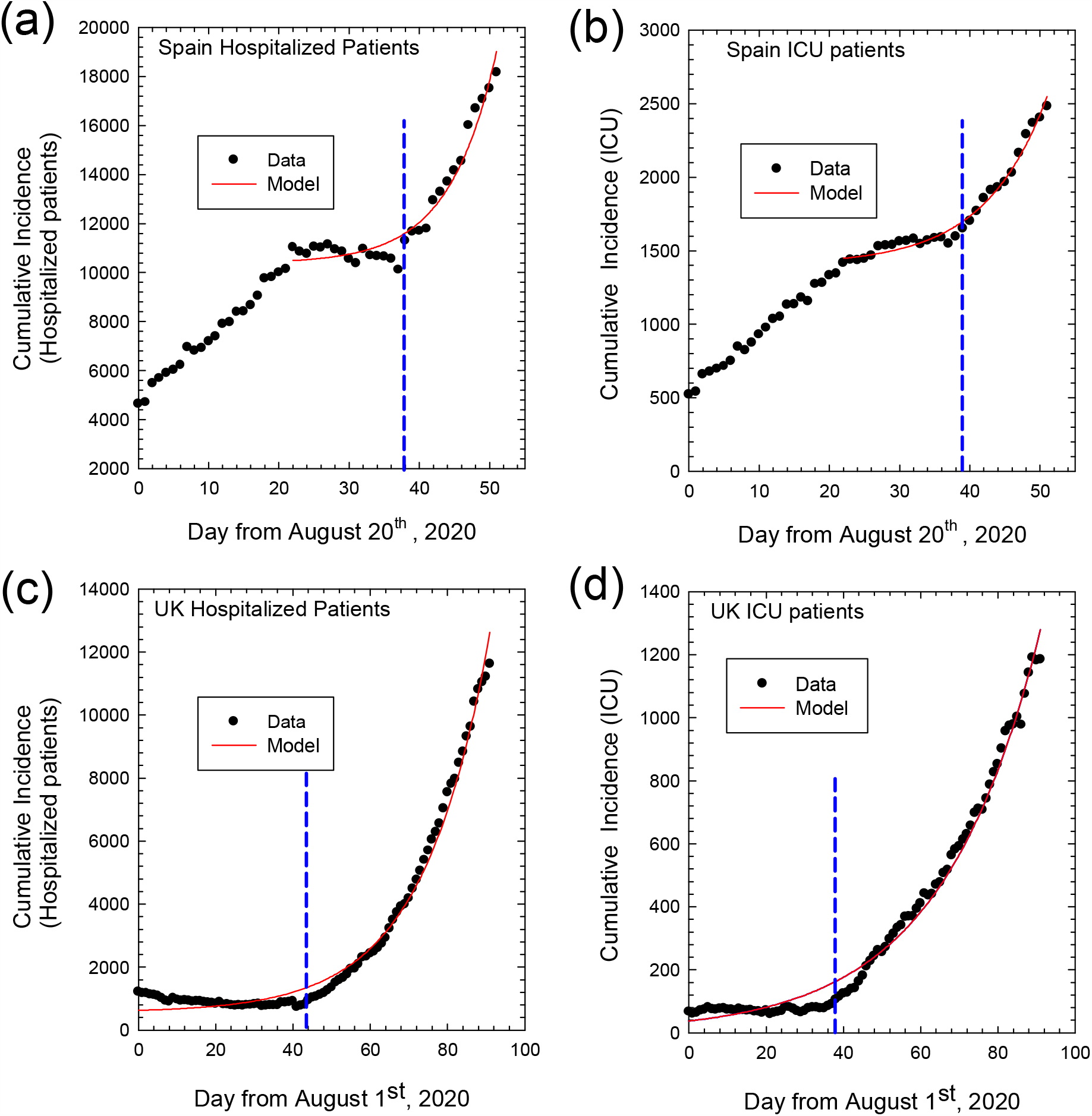
Cumulative Incidence for hospitalized and ICU patients. Cumulative incidence as numbers of total hospitalized patients (left panel) and ICU patients (right panel) from August 20^st^ to October 31^st^, 2020 for COVID-19 in Spain (a), (b) and from August 1^st^ to October 31^st^, 2020 for COVID-19 in United Kingdom (c), (d). Dots represent observed data and solid lines the fitting curves, respectively. Vertical dashed lines mark the day after which exponential regime is observed.

**Table S1:** Population and sources of data for countries considered.

|  | Resident population, Jan 1<sup>st</sup>, 2020* | Source of COVID-19 epidemic data |  |
| --- | --- | --- | --- |
|  |  | Organization | Website |
| **Austria** | 8,901,064 | Austrian Agency for Health and Food Safety (AGES) | ages.at |
| **Belgium** | 11,549,888 | Belgian Institute for Health (SCIENSANO) | sciensano.be |
| **Czech Republic** | 10,693,939 | Czech Ministry of Public Health | mzcr.cz |
| **France** | 67,098,824 | French Government | gouvernement.fr |
| **Italy** | 60,244,639 | Italian Ministry of Public Health | salute.gov.it |
| **Portugal** | 10,295,909 | Portuguese National Health System | sns.gov.pt |
| **Spain** | 47,329,981 | Spanish Ministry of Health and Social Welfare | mscbs.gob.es |
| **United Kingdom** | 67,025,542 | British Government | gov.uk |

\* Data taken from EUROSTAT (website ec.europa.eu/Eurostat)

**Table S2.** Results of epidemic parameters considered for each country

|  | Hospitalization indices |  |  |  | ICU indices |  |  |  | R<sub>ICU</sub> and related indices |  |  |
| --- | --- | --- | --- | --- | --- | --- | --- | --- | --- | --- | --- |
|  | N<sub>HP,e</sub>* | C<sub>HP</sub> | k<sub>HP</sub> | R<sup>2</sup> | N<sub>ICU,e</sub>* | C<sub>ICU</sub> | k<sub>ICU</sub> | R<sup>2</sup> | R<sub>ICU</sub> | SD | R<sup>2</sup> |
| **Austria** | 111.9 | 3.6 | 0.067 | 0.977 | 14.6 | 5.2 | 0.041 | 0.967 | 0.168 | 0.005 | 0.912 |
| **Belgium** | 226.9 | 3.2 | 0.084 | 0.993 | 66.1 | 0.3 | 0.090 | 0.996 | 0.161 | 0.002 | 0.986 |
| **Czech Republic** | 0 | 26.9 | 0.063 | 0.995 | 0 | 9.8 | 0.052 | 0.995 | 0.149 | 0.002 | 0.976 |
| **France** | 4806.7 | 11.1 | 0.082 | 0.996 | 230.5 | 76.1 | 0.041 | 0.992 | 0.178 | 0.006 | 0.819 |
| **Italy** | 1174.6 | 21.3 | 0.075 | 0.993 | 78.5 | 2.1 | 0.074 | 0.993 | 0.095 | 0.001 | 0.993 |
| **Portugal** | 320.5 | 10.7 | 0.056 | 0.993 | 32.6 | 2.9 | 0.049 | 0.990 | 0.153 | 0.003 | 0.964 |
| **Spain** | N.R. | 9.0 | 0.115 | 0.956 | N.R. | 7.0 | 0.086 | 0.962 | 0.145 | 0.003 | 0.907 |
| **United Kingdom** | 560.8 | 64.2 | 0.058 | 0.989 | 0 | 37.2 | 0.039 | 0.989 | 0.109 | 0.003 | 0.944 |

Legend: N.R = Not Reported for Spain because of the different time window selected
R<sup>2</sup> = coefficient of determination
SD= standard deviation

